# Endovascular treatment for acute ischemic stroke due to medium vessel occlusion: A systematic review with meta-analysis and trial sequential analysis

**DOI:** 10.1101/2025.04.03.25325164

**Authors:** Farsana Mustafa, Baikuntha Panigrahi, Partha Haldar, Rohit Bhatia

## Abstract

**Introduction:** A significant number of patients with acute ischemic stroke (AIS) due to medium vessel occlusion (MeVO) fail to achieve favourable functional outcomes despite the best medical treatment (BMT) currently available. The efficacy and safety of Endovascular treatment (EVT) in strokes with underlying MeVO remain uncertain, with recent randomized controlled trials (RCTs) showing no clear benefit. This meta-analysis evaluates the combined impact of these studies on efficacy and safety of EVT in MeVO patients.

**Materials and methods:** A systematic review and meta-analysis was conducted of randomized controlled trials (RCTs) in accordance with the Preferred Reporting Items for Systematic Reviews and Meta-Analyses (PRISMA) statement. A comprehensive search was conducted in five online databases, including PubMed, Scopus, EMBASE, MEDLINE and Google Scholar from inception until March 2, 2025. The primary efficacy outcome was the relative risk (RR) of excellent functional outcome, defined as mRS 0-1 at 90 days. Secondary outcomes encompassed functional independence, defined as mRS 0-2 at 90 days, alongside the primary safety outcome, which was the risk of symptomatic intracerebral hemorrhage (sICH). Other safety outcomes included serious adverse events (SAEs) and deaths.

**Results:** We included two RCTs (DISTAL and ESCAPE MeVO) involving 1,073 participants (526, EVT+BMT-arm and 547, BMT-arm). The pooled RR implied no significant difference between the two treatment arms; RR for primary efficacy outcome (mRS 0-1) was 0.95 (95% CI: 0.81-1.10) I^2^= 0%; for secondary efficacy outcome (mRS 0-2) was 0.98 (95% CI: 0.88-0.09) I^2^= 10%. The EVT+BMT-arm demonstrated a higher risk of sICH (RR: 2.39, 95% CI 1.26-4.53) I^2^= 0% and SAE (RR: 1.32, 95% CI 1.11-1.56) I^2^= 0%, while mortality at 90 days (RR: 1.29, 95% CI 0.94-1.76) I^2^=16% showed no significant difference. Trial sequential analysis revealed that for primary efficacy outcome (mRS 0-1), the z line lies in the futility zone (acquired information size AIS 96%, 1069 patients).

**Conclusion:** Our study showed that, in patients with AIS due to MeVO, EVT did not lead to better outcomes at 90 days when compared to BMT and was associated with a higher risk of sICH and SAEs compared to usual care, and this result was confirmed in a TSA analysis.

## Background and Purpose

Endovascular treatment(EVT), a highly effective treatment for ischemic stroke due to large vessel occlusion (LVO) in the internal carotid artery (ICA), M1 segment of the middle cerebral artery (MCA) and basilar artery (BA), can be considered in patients up to 24 hours after stroke onset.^1^ The meta-analysis of seven randomized controlled trials from the HERMES Collaboration, demonstrated that in patients with proximal or dominant M2 segment MCA occlusions, functional independence (mRS 0-2) at 90 days significantly improved in the EVT group compared to best medical care alone.^2^

However, a substantial proportion of patients with acute ischemic stroke (AIS) due to medium vessel occlusion (MeVO) fail to achieve favorable functional outcomes with the best medical treatment (BMT) currently available. Furthermore, approximately one third of these patients are not functionally independent 90 days after the index stroke.^3^ The prevailing European and American guidelines do not offer any recommendations or contraindications with EVT for patients with medium or distal vessel occlusion (MDVO).

Intravenous thrombolytic agents, such as alteplase and tenecteplase, have been observed to demonstrate a heightened propensity for recanalizing arteries as compared to cases of LVO. This phenomenon can be attributed to the characteristic nature of MeVO, which typically gives rise to smaller thrombi that are less fibrin-rich compared to those observed in LVO. This renders the clot more susceptible to enzymatic degradation by thrombolytic agents such as alteplase or tenecteplase.

However, a multicenter prospective study demonstrated that early recanalization following intravenous thrombolysis in the distal M1 MCA subgroup achieved in less than 50% of cases.^4^ A paucity of definitive data from prospective clinical trials exists on the efficacy and safety of EVT for the treatment of MeVO. The present systematic review and meta-analyses outlines the current evidence from RCTs conducted to evaluate the efficacy and safety of EVT for AIS among patients with MeVO. To address the existing gaps in evidence and enhance the robustness of conclusions, we also performed a trial sequential analysis (TSA). This approach allowed us to assess whether the available evidence is robust enough to inform clinical practice or if further research is warranted.

## Materials and methods

### Search Strategy

The PRISMA (Preferred Reporting Items for Systematic Reviews and Meta-Analyses) statement was employed for the purpose of reporting systematic reviews and meta-analyses in the present study.^5^ The study protocol was registered with the International Prospective Register of Systematic Reviews (PROSPERO) under the registration identification number CRD420250653970.

### Inclusion criteria

The present systematic review incorporated RCTs of patients diagnosed with AIS due to confirmed symptomatic and endovascularly treatable MDVO. The selection of patients was based on neurovascular non-invasive imaging, namely computed tomography angiography (CTA) or magnetic resonance angiography (MRA), with the caveat that the patients in question presented within 24 hours of their last known well and treated with or without intravenous thrombolysis. Exclusion criteria encompassed case reports, case series, observational studies, studies without primary efficacy outcome data, studies without primary safety outcome data and studies that did not compare endovascular therapy with no endovascular therapy in AIS due to MeVO.

### Data Extraction

A comprehensive search was conducted in five online databases by two authors independently (FM and BP): PubMed, Scopus, EMBASE, MEDLINE and Google Scholar. This search was conducted from database inception until March 2025. The MeSH terms for the search were: For Population component: “Acute stroke”, OR “Ischemic stroke” OR “cerebrovascular accident” OR “cerebral infarction” AND “thrombectomy” OR “endovascular therapy” OR “mechanical aspiration” OR “endovascular technique” OR “endovascular procedure” OR “stent retriever” AND For intervention component: “Medium Vessel Occlusion” OR “Distal Vessel Occlusion” OR “M2 Occlusion” OR “M3 Occlusion” OR “M4 occlusion” OR “A1 occlusion” “A2 Occlusion” OR “A3 Occlusion” OR “P1 Occlusion” OR “P2 Occlusion” OR “P3 Occlusion” AND For comparator component: “Best medical treatment alone” OR “Intravascular thrombolysis” AND For the outcome component: “Excellent functional outcome” OR “ Best functional outcome” OR “Symptomatic Intracranial hemorrhage” OR “ Deaths” AND “Randomized Controlled Trial” OR “RCT” with filters to include only RCTs. In the present meta-analysis, Covidence software (https://www.covidence.org/) was utilized for the systematic study selection, duplication removal and data extraction, thereby ensuring a standardized and efficient data management process. Titles and abstracts were independently evaluated by two authors (F.M. and B.P.) to ascertain their inclusion and to determine whether the full text should be screened. Any discrepancies were resolved by R.B. Full texts were retrieved for further consideration with regard to inclusion in the study. However, the individual patient data sets of the included studies were not available for review.

### Outcomes

The primary efficacy outcome extracted from the studies was defined as the relative risk of achieving an excellent functional outcome, which was categorised as a modified Rankin Scale (mRS) score of 0–1 at 90 days. The secondary outcome encompassed functional independence, measured by an mRS score of 0–2 at 90 days. The safety outcomes encompassed symptomatic Intracranial hemorrhage (sICH), death from any cause at 90 days and any serious adverse events within 90 days post-intervention.

### Assessment of risk of bias

Following the selection of relevant studies, the data was extracted onto MS Excel by two authors independently (FM and BP). The information extracted comprised the first author, the year of publication, the number of patients included in each study, mono or multi-centric study, the baseline characteristics of patients included in each study, mRS at 90 days, sICH, death from any cause and any serious adverse events. Any discrepancy between the two authors was resolved in consultation with a third author (RB). The Cochrane Risk of Bias (ROB2) tool was utilized to evaluate the risk of bias in RCTs.

### Statistical Analysis

Statistical analyses were conducted using the R Project for Statistical Computing (https://www.r-project.org/). A common effects model (fixed effects model) was employed due to minimal heterogeneity (I^2^ = 0%, τ^2^ = 0, p = 0.77), indicating negligible variation across studies. Risk ratios (RR) with corresponding 95% confidence intervals (CI) were utilized to generate the forest plot. Statistical heterogeneity was assessed using Higgins I^2^ statistic, with values of I^2^ = 0% suggesting no detectable between-study variability. The classification of study-level estimates was determined by the I^2^ value, with estimates categorized as heterogeneous if the I^2^ value exceeded 50%. An I^2^ value between 50% and 75% was considered indicative of substantial heterogeneity, while a value greater than 75% was regarded as considerable heterogeneity. A subgroup analysis was performed based on occlusion location (M2, M3, ACA, PCA), intravenous thrombolysis status (EVT with IVT vs EVT alone), and time to reperfusion (4.5 hours vs. 6-24 hours). A random-effects model was applied for pooled estimates, and meta-regression was used to evaluate effect modification by these covariates. Since this meta-analysis included a limited number of eligible studies, resulting in a relatively small number of overall randomized participants, it was necessary to assess whether the analysis had sufficient statistical power to reliably evaluate the intervention effects. Consequently, for the primary efficacy outcome, a Trial Sequential Analysis (TSA) was also conducted, in which trials were included in chronological order and analysed as interim analyses relative to the required number of randomized participants. TSA accounted for the required information size (RIS) and established monitoring boundaries to control for Type I (false positive, alpha boundary) and Type II (false negative, beta boundary) errors. A TSA-adjusted 95% confidence interval and an adjusted threshold for statistical significance, lower than the conventional p < 0.05, were reported if the required information size (RIS) had not been reached. If the line representing the cumulative z score fails to cross the alpha boundary, we conclude that the existing studies do not provide sufficient evidence to show a significant difference relative to RIS; if the z line enters the beta boundaries, contingent upon achieving RIS, we can declare futility. We also assessed certainty of the evidence using the GRADE pro GDT tool.

## Results

### Study selection

A total of 1115 studies were identified, of which 312 duplicates were removed. 803 studies underwent title and abstract screening and 799 studies were found irrelevant. Four studies underwent full text screening (ESCAPE MeVO,^6^ DISTAL^7^ DUSK study^8^ and DISCOUNT study^9^). The DUSK study was a retrospective analysis of prospectively collected data, and the DISCOUNT study was excluded as it was a protocol. A total of two randomized control trials (DISTAL and ESCAPE Mevo) comprising 1,073 patients met the predefined inclusion and exclusion criteria. Of these patients, 526 were in the EVT arm and 547 were in the best medical treatment (BMT) arm. The details are depicted in the PRISMA flow chart (Figure 1).

**Figure 1:**
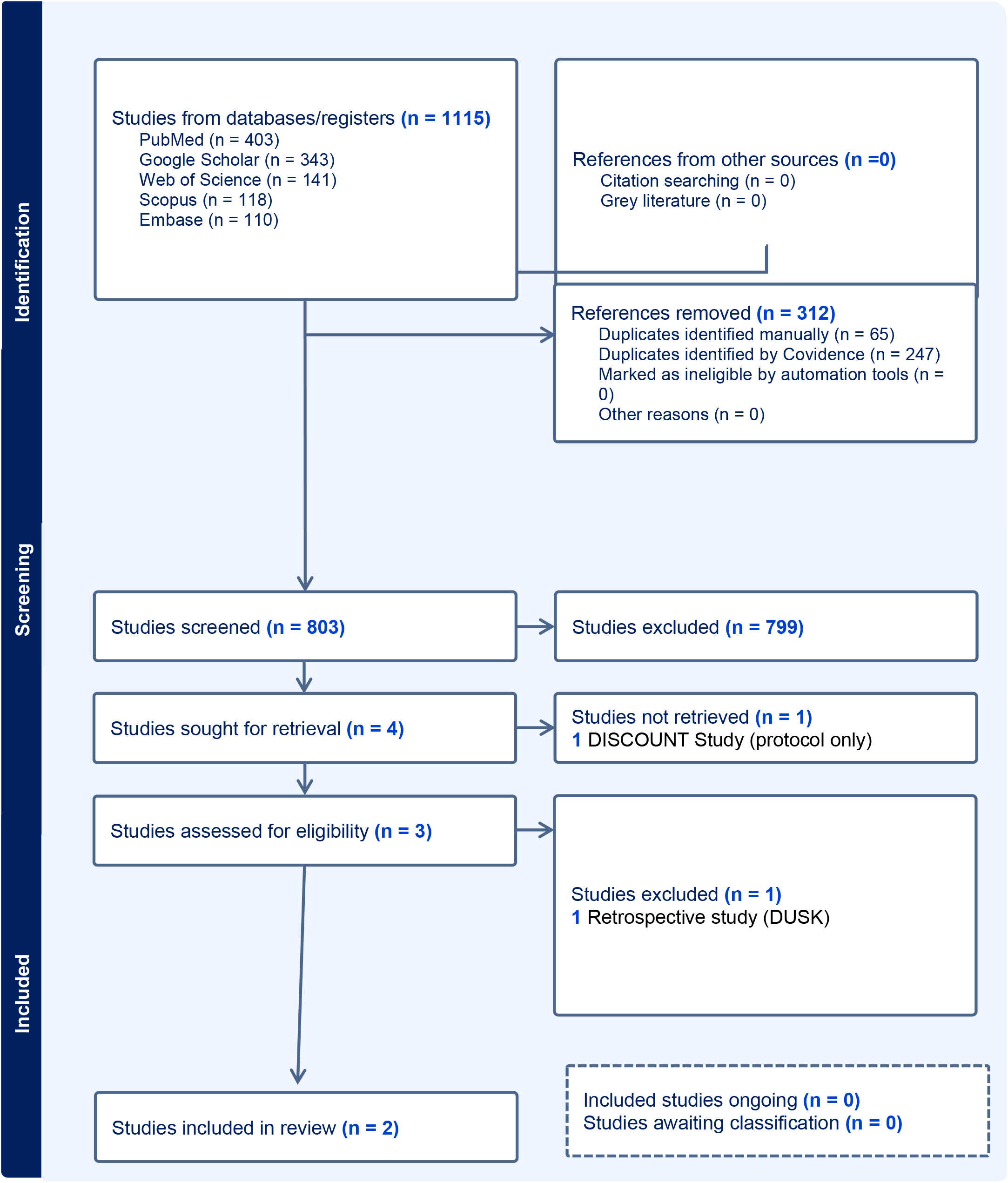
PRISMA flow chart

### Study characteristics

The baseline characteristics of the patients included in the two studies are shown in Table 1. No significant differences in baseline characteristics were observed between the EVT and BMT arms in both studies. The median National Institute of Health Stroke Scale (NIHSS) at admission was found to be marginally higher in the ESCAPE MeVO trial group compared to the DISTAL trial. The ESCAPE MeVO trial made a specific exclusion of occlusions of the M1 segment, A1 segment, and P1 segment, as well as M4 occlusions. This is in contrast to the DISTAL trial, which included a small proportion of M4 occlusions and certain cases of A1 and P1 occlusions. The prevalence of M2 occlusions was marginally higher in DISTAL EVT, while ESCAPE MeVO was observed to have a higher proportion of M3 occlusions. The DISTAL trial had a higher number of patients who received intravenous thrombolysis (IVT) compared to the ESCAPE MeVO trial (355 vs. 305 patients, respectively).

### Risk of bias assessment across studies

Both the trials included in the systematic review are having low risk of bias in the randomization domain. Concerns were raised regarding the potential for bias due to deviations from the intended interventions (i.e., adherence to the intervention). This was due to the participants and their caregivers or intervention providers being aware of the interventions. The missing outcome data were addressed through the implementation of imputation methods (Table2). One trial utilized multiple imputation by chained equations (MICE), while the other employed a last score carried forward approach, imputing a score of 4 in instances where both day 30 and day 90 scores were missing but the patient was alive at 90 days. These methodologies were employed to minimize bias and ensure the integrity of the data. In both trials, the blinded outcome assessors were successful in minimizing detection bias. The primary outcome analysis was conducted in accordance with a pre-specified plan, thereby ensuring consistency. Overall, the open label design introduces some bias, but randomization and blinded outcome assessment help mitigate its impact by ensuring balanced groups and objective outcome evaluation (Figure 2).

**Figure 2:**
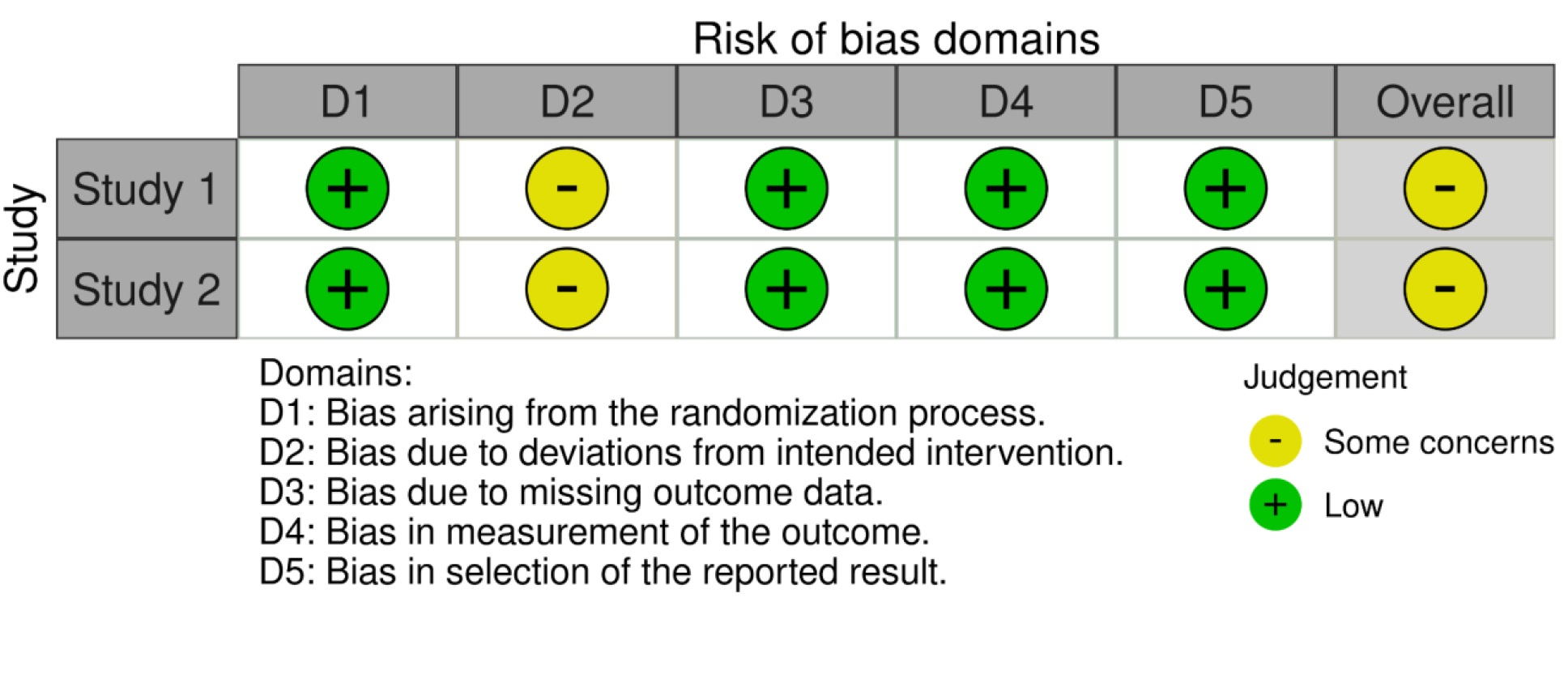
Traffic plot illustrating the risk of bias across different domains for each study

## Outcomes

### Excellent functional outcome (mRS 0-1) at 90 days

The primary efficacy outcome, defined as excellent functional outcome (mRS 0–1 at 90 days), was achieved in 200 out of 526 patients (38%) in the EVT plus BMT arm, and 219 out of 543 patients (40.3%) in the BMT arm (Table 3). The pooled relative risk (RR), calculated using the common effects model was 0.95 (95% CI: 0.81-1.10), [95% TSA−adjusted CI: 0.79-1.13] indicating no significant difference between the two treatment arms. The I^2^ test for heterogeneity was 0%, demonstrating consistency across studies (Figure 3A). The TSA showed that with an achieved information size (AIS) of 1069 (96%), the line representing the cumulative z-score failed to cross the alpha boundary, implying that the existing studies do not provide sufficient evidence to show a significant difference between EVT-plus-BMT arm vs BMT alone; additionally, the cumulative z-score line has already entered the beta boundaries in the futility zone (Figure 4).

**Figure 3:**
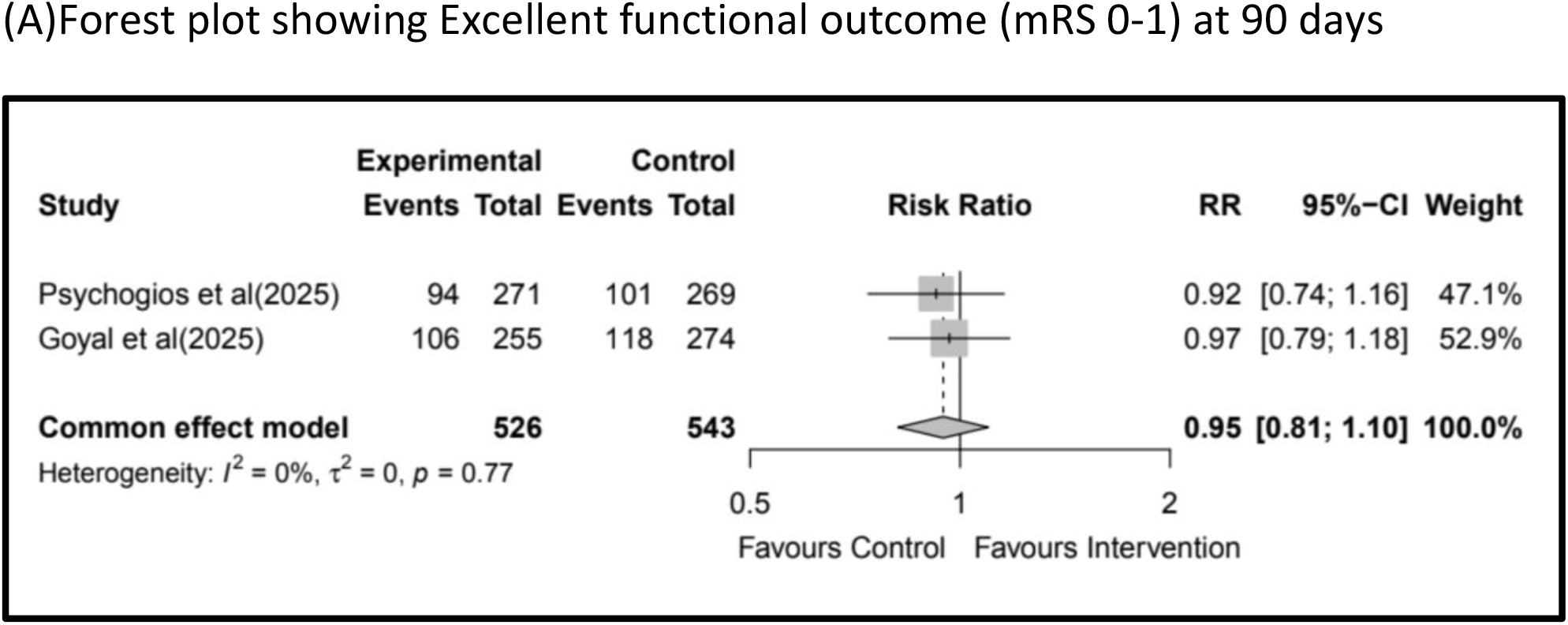
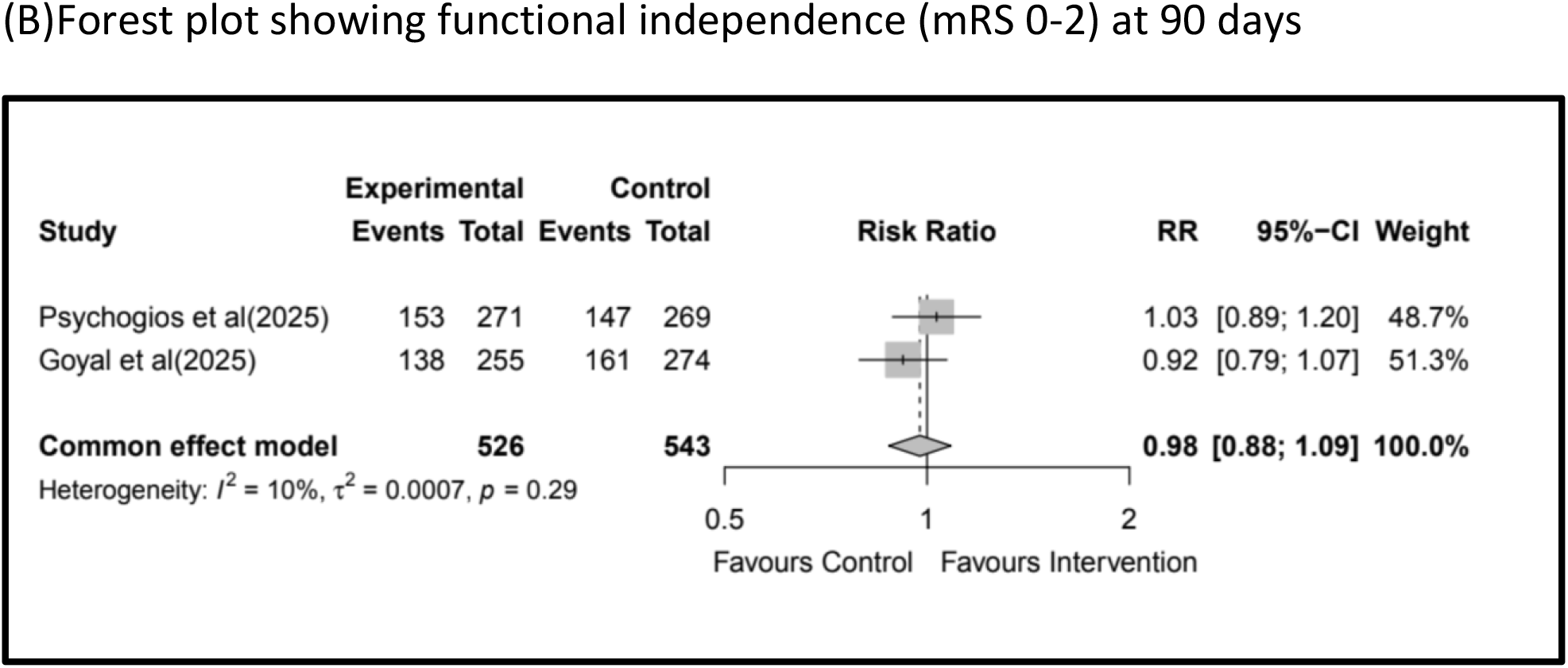
Forest plot of the efficacy outcomes (A) Forest Plot for mRS 0-1; (B) Forest Plot for mRS 0-2

**Figure 4:**
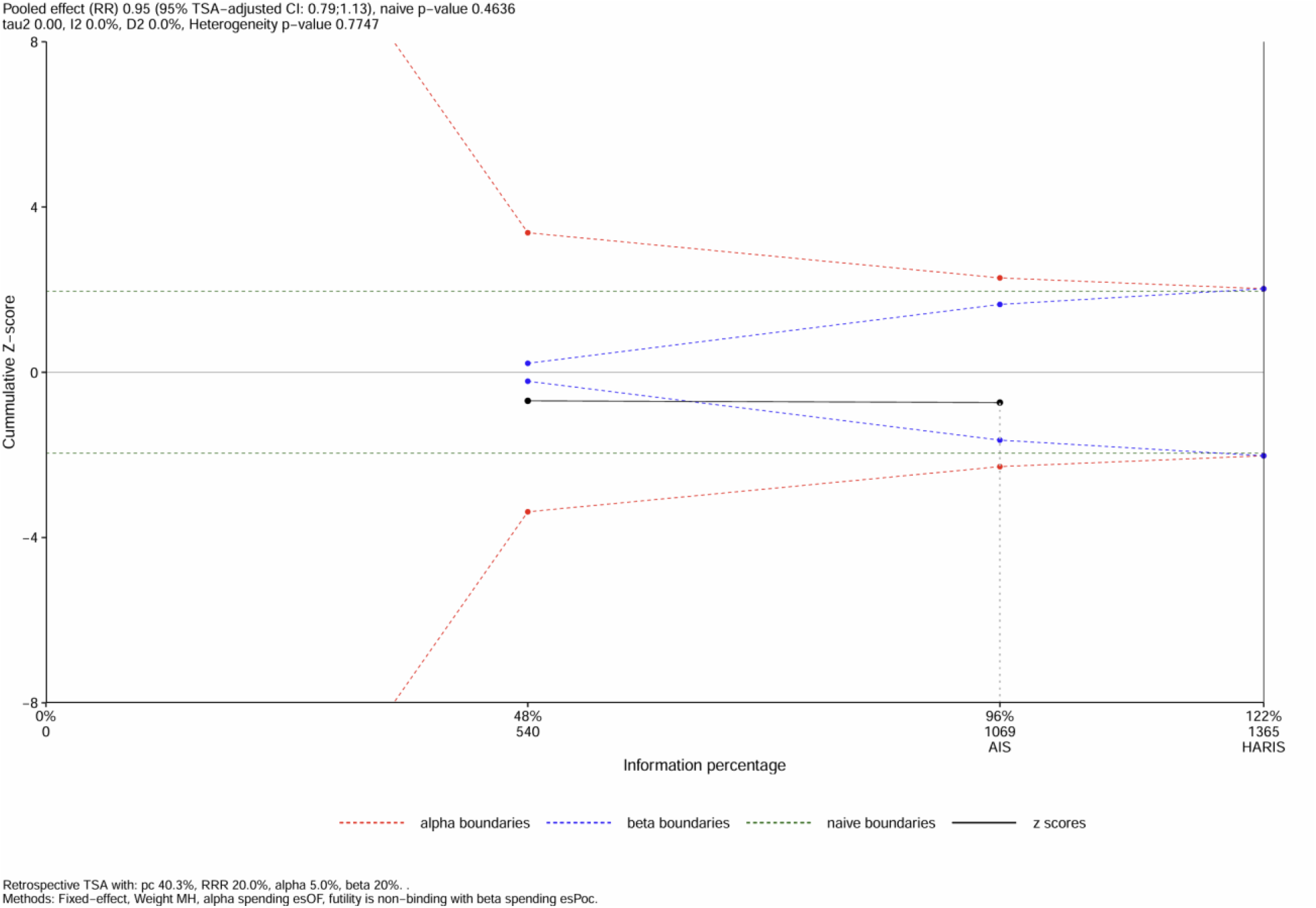
Trial sequential analysis plot evaluating the cumulative evidence from included studies on a pooled relative risk effect. The x-axis represents the cumulative information size (number of participants/events, in percentage of the required information size), while the y-axis shows the cumulative Z-score for the meta-analysis

### Functional independence (mRS 0-2) at 90 days

The secondary efficacy outcome, defined as functional independence (mRS 0–2 at 90 days post-intervention), was achieved in 291 out of 526 patients (55.32%) in the EVT plus BMT arm, and 308 out of 543 patients (57%) in the BMT arm (Table 3). The pooled RR using the common effects model was 0.98 (95% CI: 0.88,1.09), indicating no significant difference between the two treatment arms (Figure 3B). The heterogeneity for this outcome was low (I^2^ = 10%), indicating minimal variability between studies, suggesting that the results were consistent across trials, and the pooled estimate is reliable for interpretation (Figure 3B).

### Safety outcomes

#### Symptomatic Intracranial hemorrhage (s ICH) within 24 hours

The primary safety outcome, defined as symptomatic intracranial hemorrhage (sICH) within 24 hours, occurred in 30 out of 526 patients (5.7%) in the EVT plus BMT arm and 13 out of 546 patients (2.4%) in the BMT arm (Table 3). The pooled RR, calculated using the common effects model, was 2.39 (95% CI: 1.26-4.53), indicating a significantly higher risk of sICH in the EVT plus BMT arm. Additionally, the I^2^ test for heterogeneity was 0%, demonstrating consistency across studies (Figure 5A).

**Figure 5:**
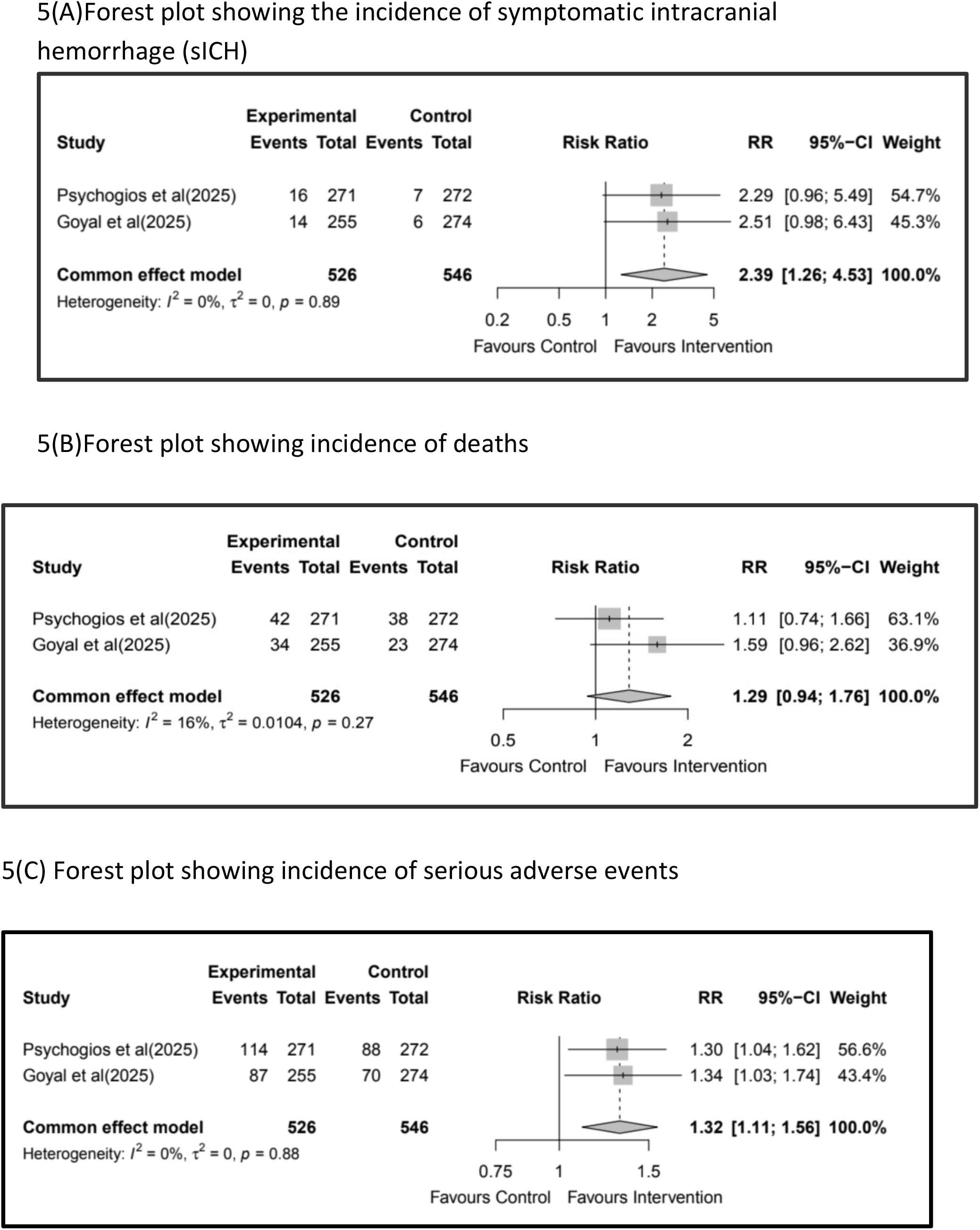
Safety outcomes: (A) Forest Plot of sICH (B) Forest Plot of any serious adverse events (C)Forest plot of all deaths

#### Deaths from any cause at 90 days

The secondary safety outcome, defined as death from any cause at 90 days, occurred in 76 out of 526 patients (14.4%) in the EVT plus BMT arm and 61 out of 546 patients (11.2%) in the BMT arm (Table 3). The pooled RR, calculated using the common effects model, was 1.29 (95% CI: 0.94–1.76), indicating no significant difference in mortality between the two treatment arms. Additionally, the I^2^ test for heterogeneity was 16%, suggesting low variability between studies and moderate consistency in findings (Figure 5B).

#### Serious adverse effects within 90 days

The secondary safety outcome, defined as serious adverse events (SAEs) within 90 days, occurred in 201 out of 526 patients (38.2%) in the EVT plus BMT arm and 158 out of 546 patients (28.9%) in the BMT arm (Table 3). The pooled RR, calculated using the common effects model, was 1.32 (95% CI: 1.11–1.56), indicating a significantly higher incidence of serious adverse events in the EVT plus BMT arm. Additionally, the I^2^ test for heterogeneity was 0%, suggesting no significant variability between studies and high consistency in findings (Figure 5C).

## Discussion

Our analysis suggested that among patients with AIS due to MeVO who presented within 24hrs of last being seen normal and had evidence of salvageable tissue on baseline imaging EVT plus BMT did not significantly improve the probability of achieving an excellent functional outcome at 90 days compared to BMT alone. The findings of this review also suggested that there is no statistically significant improvement in functional independence when EVT is used in conjunction with BMT, as opposed to BMT alone in those patients. The TSA further supported the non-significant effect, and also futility of EVT-plus-BMT arm bs BMT alone, with a robust achieved information size (96%). We also reported that the EVT plus BMT arm had a higher risk of sICH and SAE, while mortality at 90 days showed no significant difference. A low level of heterogeneity while pooling the results indicated consistency across the studies.

We had excluded the DISCOUNT trial (NCT05030142) because, the limited results (trial stopped following first interim analysis) available from this trial as a conference abstract did not have data on the primary outcome (mRS-0-1). It included AIS patients with MeVO presenting within 24 hours of last being seen well, with compatible advanced imaging and an NIHSS ≥ 5 or disabling aphasia. The trial randomized 163 patients with occlusions distal to the mid-insula for the MCA and all anterior (A1–A3) and posterior cerebral (P1–P3) artery occlusions.^9^ The primary outcome, mRS 0–2 at 90 days, occurred in 60% of patients treated with EVT plus BMT versus 77% in the BMT-alone group (OR 0.42, 95% CI 0.20– 0.88, after multiple imputations).

The results of our meta-analysis of RCTs are significantly different from the previous post-hoc analyses of randomized and non-randomized studies showing a benefit of EVT in MeVO. The HERMES collaboration meta-analysis employed individual patient-level data from multiple major RCTs of EVT in LVO stroke, thus facilitating detailed subgroup analyses, including patients with M2 segment MCA occlusions. This demonstrated that EVT in patients with M2 segment MCA occlusions significantly improved functional outcomes (mRS 0–2: 58.2% vs. 39.7%; OR 2.39, 95% CI 1.08–5.28), with efficacy comparable to that seen in proximal occlusions.^2^ This difference could be due to multiple factors.

First, the reperfusion rates in these trials were significantly lower than LVOs. For example, in the DISTAL trial, successful reperfusion was achieved in 71.7% of patients in the EVT plus BMT group, while in the ESCAPE MeVO trial, it was observed in 75.1% of cases, likely influenced by technical challenges.^6,7^ Despite the technical differences between the MeVO-eTICI and eTICI scales, previous LVO trials have reported successful reperfusion rates of 85% or higher.^10,11^ This low incidence of successful reperfusion rates may have been a contributing factor to the neutral findings of EVT observed in the review. Serious adverse events and symptomatic intracranial hemorrhage were more prevalent in the intervention group than in the usual care group.^6^ These may have contributed cumulatively to the neutral findings in the primary outcome.

Secondly, in both trials, the median workflow time for EVT was longer than in previous LVO trials, which may have reduced the procedure’s benefit by limiting the amount of salvageable tissue. For example, in the DISTAL trial, the median time from imaging to arterial puncture was 70 minutes, thus exceeding the target of 60 minutes, despite efforts to expedite EVT delivery. This delay was primarily due to a longer interval between imaging and randomization, which was attributed to challenges in detecting distal and medium vessel occlusions, as well as the time required to obtain patient consent. The workflow in the ESCAPE-MeVO trial also revealed a longer median time from onset to recanalization of 359 minutes, in comparison to the 249 minutes observed in the ESCAPE trial.^12^ The time delay in the ESCAPE-MeVO trial was partly attributed to the time required to arrange general anesthesia and technical challenges in achieving reperfusion in distal arteries.

Additionally, distal arteries are end arteries without collaterals, emphasizing the critical importance of timely treatment in MeVO strokes, as they have less compensatory capacity compared to LVO strokes.^14^ In the ESCAPE-MeVO trial, a minority of patients with MeVO (39 out of 255) in the EVT arm experienced spontaneous recanalization between noninvasive imaging and the first intracranial angiographic assessment, thereby eliminating the need for thrombectomy despite undergoing angiography. In the majority of cases (32 out of 39), this recanalization was attributed to intravascular thrombolysis, thereby highlighting that spontaneous thrombus resolution can occur in MeVO following thrombolysis.^6^

The anatomical complexity of the vessels, characterized by their reduced diameter and distal location, poses a significant challenge to the efficacy of currently employed devices.^13^ These devices have been primarily designed for LVOs, and consequently, their application in the context of MeVO strokes may result in suboptimal perfusion outcomes. Additionally, the operator’s experience in performing EVT procedures can also influence the success rates of such interventions. The fragility and smaller size of these arteries render them more susceptible to injury during the procedure. Consequently, these factors influence the safety outcomes of sICH and SAEs in the intervention subgroup. A refined patient selection process may be necessary to identify those who are most likely to benefit while minimizing harm in such case scenarios. Improved patient selection is essential for optimizing outcomes. First, establishing a standardized definition for M2 segment and subsegment occlusions would create a more homogeneous patient group. Second, raising the NIHSS threshold might help identify patients most likely to benefit from EVT. For example, subgroup analysis from the DISTAL trial showed a trend toward better outcomes with MT as NIHSS increased. This aligns with a population-based study on M2 occlusions, which identified an NIHSS cut-off of 9 points as predictive of poor outcomes, with 78% of patients scoring >9 having unfavorable results compared to 22% of those scoring <9.^14^Advancements in EVT devices and techniques should also parallel improved patient selection. In the ESCAPE-MeVO trial, only 75% of recanalization were successful (adapted mTICI 2b-3 scores). Achieving complete recanalization on the first pass is crucial, as the first-pass effect is linked to better outcomes. Additionally, subgroup analysis from the DISTAL trial suggested fewer hemorrhagic complications with aspiration catheters compared to stent retrievers or a combination of both. It is also imperative to optimize workflow efficiency, improve imaging to treatment time, and streamline the decision making process in order to mitigate these delays and improve EVT outcomes in MeVO patients.

The principal limitation of this study is the paucity of randomized controlled trials published on this research question to date. Moreover, the lack of access to individual patient data restricts the ability to perform subgroup analyses, which could have identified specific patient populations more likely to benefit from EVT in MeVOs.

## Conclusions

While EVT remains a promising intervention for stroke, its role in MeVO requires further investigation to determine optimal strategies for improving patient outcomes while mitigating risks associated with the procedure.

## Data Availability

All data produced in the present work are contained in the manuscript

## Sources of funding

No funding sources to disclose

## Disclosures

None of the authors have any conflict of interest or competing interest

## Notes

### Competing Interest Statement

The authors have declared no competing interest.

### Funding Statement

This study did not receive any funding

